# Allelic expression imbalance in articular cartilage and subchondral bone refined genome-wide association signals in osteoarthritis

**DOI:** 10.1101/2022.04.07.22273552

**Authors:** Rodrigo Coutinho de Almeida, Margo Tuerlings, Yolande Ramos, Wouter Den Hollander, Eka Suchiman, Nico Lakenberg, Rob (RGHH) Nelissen, Hailiang Mei, Ingrid Meulenbelt

**Affiliations:** Department of Biomedical Data Sciences, Section Molecular Epidemiology, Leiden University Medical Center, Leiden, The Netherlands; Department of Orthopaedics Leiden University Medical Center, Leiden, The Netherlands; Sequencing Analysis Support Core, Dept. of Biomedical Data Sciences, Leiden University Medical Center, Leiden, The Netherlands

## Abstract

Osteoarthritis (OA) is an age-related joint disease with a strong and complex genetic component. Genome-wide association studies (GWAS) discovered a large number of genomic regions associated with OA. Nevertheless, to link associated genetic variants affecting the expression of OA-risk genes in relevant tissues remains a challenge. Here, we showed an unbiased approach to identify transcript single nucleotide polymorphisms (SNPs) of OA risk loci by allelic expression imbalance (AEI). We used RNA sequencing of articular cartilage (N = 65) and subchondral bone (N= 24) from OA patients. AEI was determined for all genes present in the 100 regions reported by GWAS catalog. The count fraction of the alternative allele (φ) was calculated for each heterozygous individual with the risk-SNP or with the SNP in linkage disequilibrium (LD) with it. Furthermore, a meta-analysis was performed to generate a meta-φ (null hypothesis median φ=0.49) and P-value for each SNP. We identified 30 transcript SNPs subject to AEI (28 in cartilage and 2 in subchondral bone). Notably, 10 transcript SNPs were located in genes not previously reported in the GWAS catalogue, including two long intergenic non-coding RNAs (lincRNAs), *MALAT1* (meta-φ=0.54, FDR=1.7×10^−4^) and *ILF3-DT (*meta-φ=*0*.*6, FDR=1*.*75×10*^*-5*^). Moreover, 14 drugs were interacting with 7 genes displaying AEI, of which 7 drugs has been already approved. By prioritizing proxy transcript SNPs that mark AEI in cartilage and/or subchondral bone at loci harboring GWAS signals, we present an unbiased approach to identify the most likely functional OA risk-SNP and gene. We identified 10 new potential OA risk genes ready for further, translation towards underlying biological mechanisms.

## Introduction

Osteoarthritis (OA) is an age-related, joint disease, characterized by progressive heterogeneous changes in articular cartilage and subchondral bone. Over 80% of OA patients have limitations in movement and 25% inhibition in major daily activities of life (1,2). Up until now, there is no disease modifying treatment except for costly total joint replacement surgery at end-stage disease. As a result, OA puts a high social and economic burden on society (3). OA has a considerable and complex genetic component (4) and well-powered genetic studies are accumulating at high rate, highlighting strong OA risk genes and associated underlying disease pathways. By addressing functionality of identified OA risk alleles, such as for example for *COLGAT2* (5) and *MGP* (6), it was confirmed that OA risk single nucleotide polymorphisms (SNPs) frequently modulated OA pathology due to subtle altered transcription of a positional gene *in cis*, as such imposing a persistent negative influence on joint tissue homeostasis throughout life (6). Targeted testing of allele imbalanced expression of OA risk genes is, however, cumbersome (7–9). To address the full potential of biologically relevant DNA variants in cartilage, we previously captured allelic expression imbalance (AEI) at a genome wide scale, by virtue of a relatively small transcriptomics (RNA sequencing) dataset of paired preserved and lesioned OA cartilage (10). This study presented, however, a database of AEI SNPs in cartilage for researcher to probe their gene of interest without providing the linkage disequilibrium (LD) structure relative to the OA risk SNPs. Moreover, genome wide association studies (GWAS) in OA with ever increasing sample sizes have resulted in increasing number of OA risk SNPs with 100 currently mapped loci associating to OA related phenotypes (7). Finally, given the pathways in which the genes located in these loci act, the AEI analyses should be extended to other OA relevant tissues and particularly subchondral bone as was recognized by recently studies (5,8,9)

In the current study we used a relatively large RNA-sequencing dataset of OA cartilage and subchondral bone to capture AEI of transcript SNPs that tag OA risk SNPs recently reported by GWAS, as such ready for further study towards underlying biological mechanisms e.g. in human 3D *in vitro* model systems incorporating bone and/or cartilage tissue units.

## Results

### Identification of Allelic Expression Imbalance of transcript SNPs tagged by the OA risk SNPs

We initially performed a look-up on the expression levels in our previously published cartilage and subchondral bone datasets (11,12) for genes located at all 100 OA risk loci reported by GWAS catalog (7,13) residing within a 1Mb window. To obtain relevant information on the functional effect of the identified OA risk alleles on expression of positional genes, we next prioritized on positional transcript SNPs that were in LD (r^2^> 0.6) to the OA risk SNP and subject to AEI.

We identified 28 transcript SNPs subject to AEI in cartilage of positional genes in LD with the most associated risk SNP reported on OA GWAS (**Figure 1**; **Supplementary Table 1**). Notable among cartilage, we identified 10 AEI SNPs located in genes not previously highlighted as possible OA genes candidates according to GWAS catalogue (**Table 2**), including two long intergenic non-coding RNAs (lincRNAs) such as, *MALAT1* (AEI meta-φ=0.54, FDR=1.7×10^−4^) and *ILF3-DT* (meta-φ=0.6, FDR=1.7×10^−5^) (**Figure 2A-B**). Moreover, we identified two SNPs subject to AEI in subchondral bone of OA patients affecting the expression of three GWAS reported genes, the HLA genes *HLA-DPA1* and *HLA-DPB1 (*meta-φ=0.76, FDR=0.0033; **Figure 2C**) and MGP (meta-φ=0.33, FDR = 1.1 x10^−19^) (**Supplementary Table 1**).

**Figure 1:**
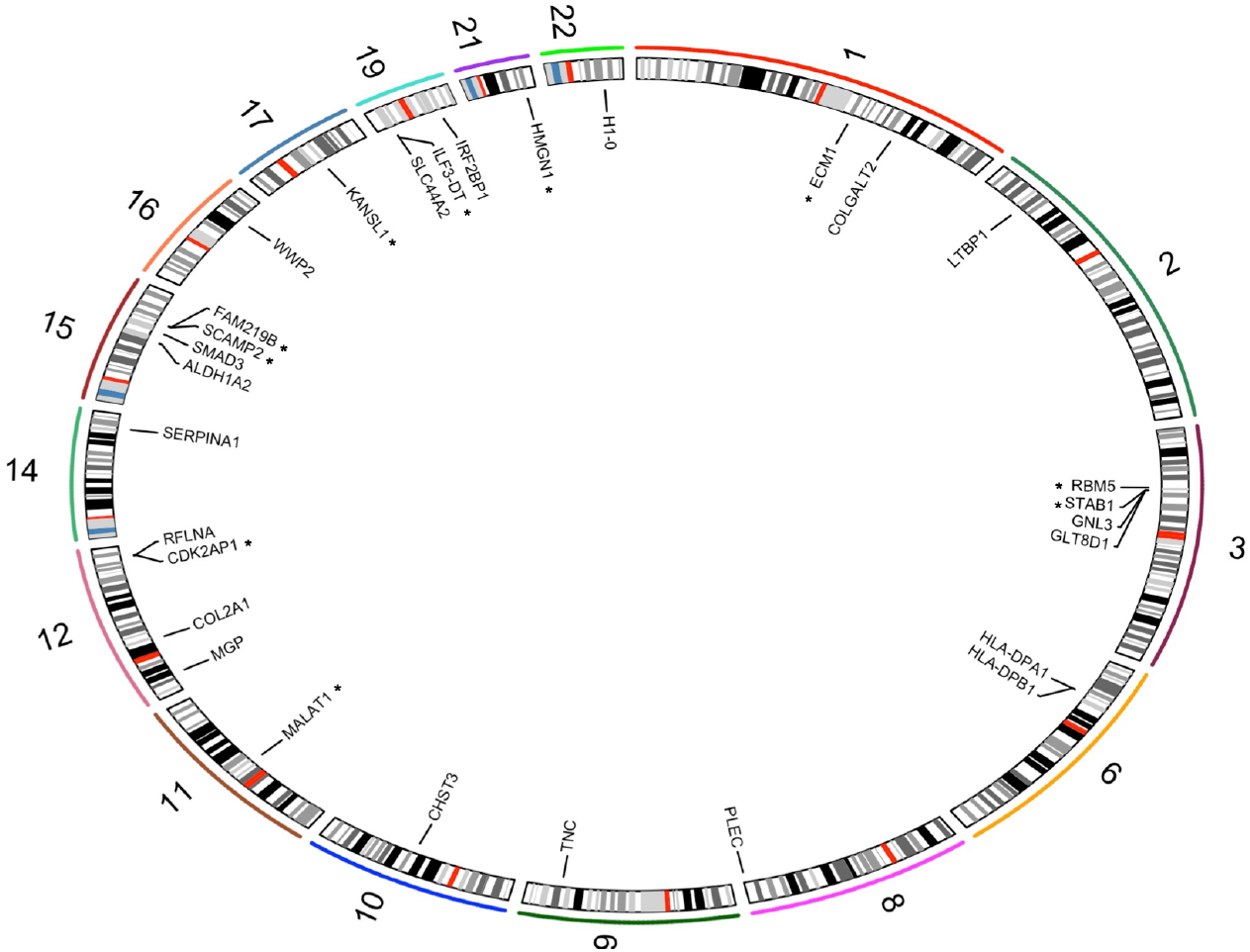
Circos plot showing all genes with allelic expression imbalance in OA across the genome. Stars represents genes that were not previously reported by GWAS catalog.

**Figure 2:**
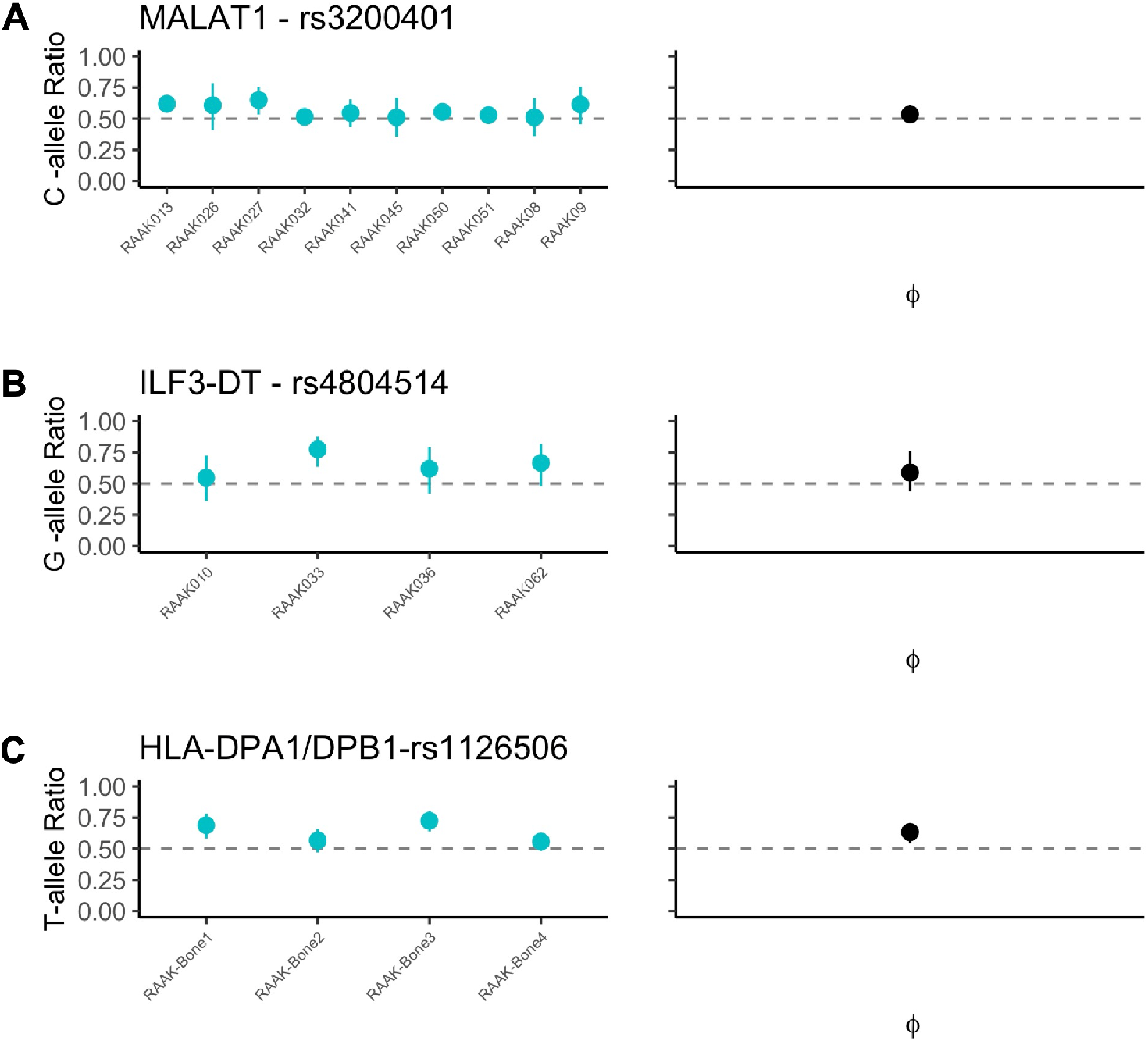
Allelic expression imbalance of two lincRNAs in articular cartilage and HLA genes in subchondral bone. (A) SNP rs3200401 showing increasing expression of lincRNA MALAT1 of the risk allele C in OA cartilage (meta-φ= 0.54). (C) LincRNA ILF3-DT with risk allele G displaying increasing expression in OA cartilage (meta-φ = 0.6). (D) HLA-DPA1/DPB1 with risk allele T increasing expression in OA subchondral bone (meta-φ = 0.76).

### AEI genes differentially expressed between preserved and lesioned in cartilage and subchondral bone

To further strengthen that these genes are likely involved in OA pathophysiology, we lookup for differential expression between preserved and lesioned articular cartilage (N=35 paired) and subchondral bone (N=24) in our previously published datasets (11,12). We found nine AEI genes differentially expressed in cartilage and two in subchondral bone (**Table 3**), including *TNC* gene (meta-φ=0.47, FDR=2.08×10^−7^, FC=1.4) which showed a downregulation AEI for the risk allele but upregulated in lesioned cartilage, suggesting that its expression in lesioned cartilage is merely a beneficial response to the occurring OA pathophysiological process (**Figure 3A**). On the other hand, the *COLGALT2* gene was differentialy expressed between preserved and lesioned cartilage and with an upregulated AEI effect in cartilage (meta-φ=0.54, FDR = 0.00132, FC=1.4; **Figure 3B**), suggesting that a lower expression in lesioned cartilage could be potentially beneficial. Additionally, *SLC44A2* gene was differentially expressed in subchondral bone and in cartilage (**Table 3**), however with AEI was exclusively in cartilage (meta-φ=0.65, FDR=1.13×10^−11^, **Figure 3C**), indicating that lower expression of SLC44A2 in cartilage is conferring risk to OA likely by an aberrant cartilage specific transcription factor.

**Figure 3:**
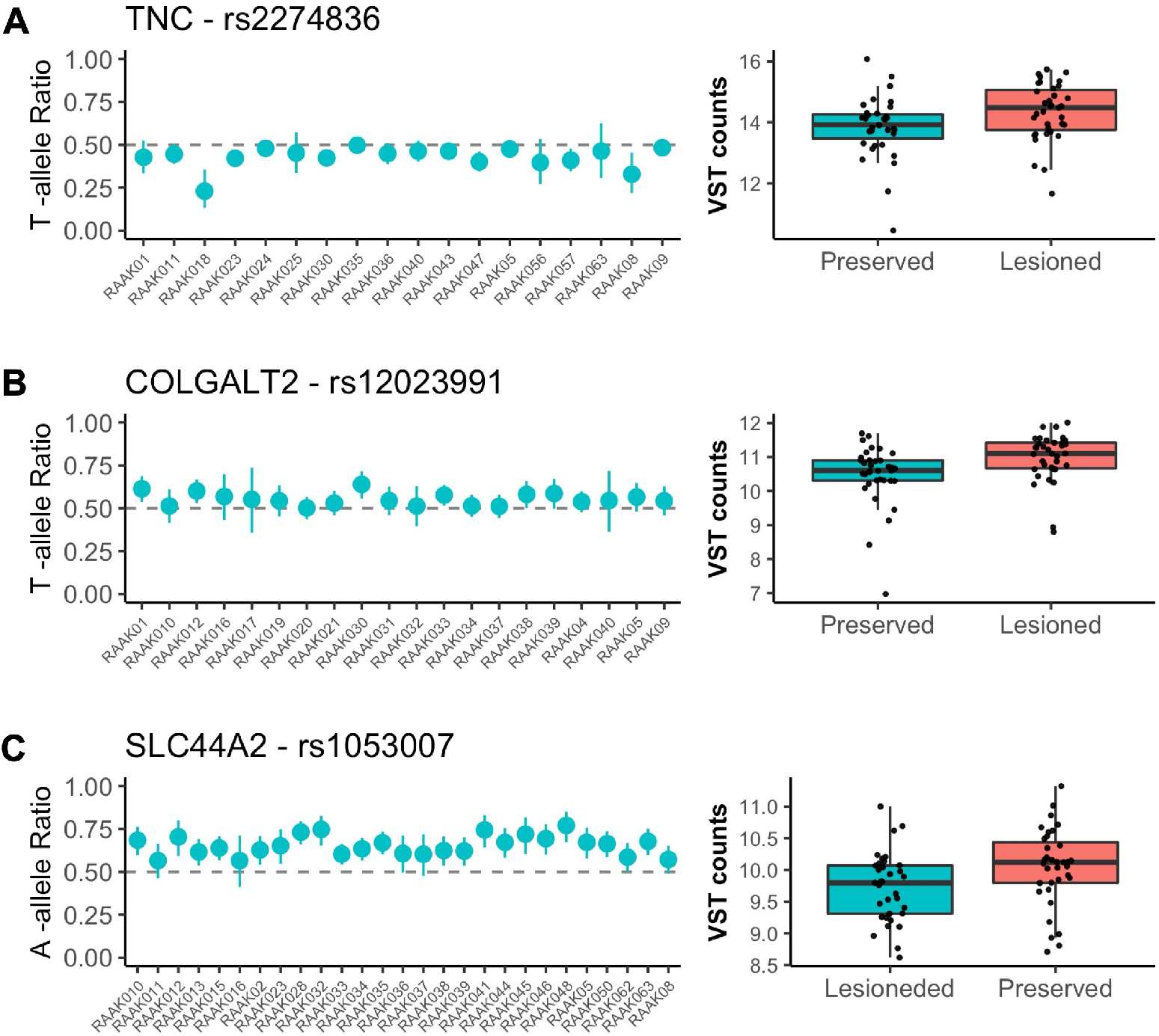
Genes with allelic expression imbalance (AEI) also showing differentially expressed between preserved and lesioned cartilage. (A) TNC upregulated in lesioned cartilage and with AEI downregulated effect (meta-φ = 0.47); (B) *COLGALT2*. (meta-φ= 0.54); (C) *SLC44A2* (meta-φ = 0.65).

### AEI genes interacting with 14 drugs

By assessing effects of identified OA risk alleles on expression of positional genes, a reliable inference can be made on potential druggable targets that could counteract this aberrant gene expression. Therefore, we next combining OA risk alleles subject to AEI to known drugs and their targets on all 30 AEI genes. In total 7 genes showed interaction with at least one drug (DGBI IG score > 0.5). Out of 15,000 drugs present on the DGBI database, 14 drugs showed 2.1-fold significant enrichment (Fisher’s exact test p-value = 0.044) interaction with an AEI gene with a mode of action consistent with the direction of the risk allele expression (**Figure 4; Supp Table 2**). In addition, seven drugs have been already approved by federal agencies.

**Figure 4:**
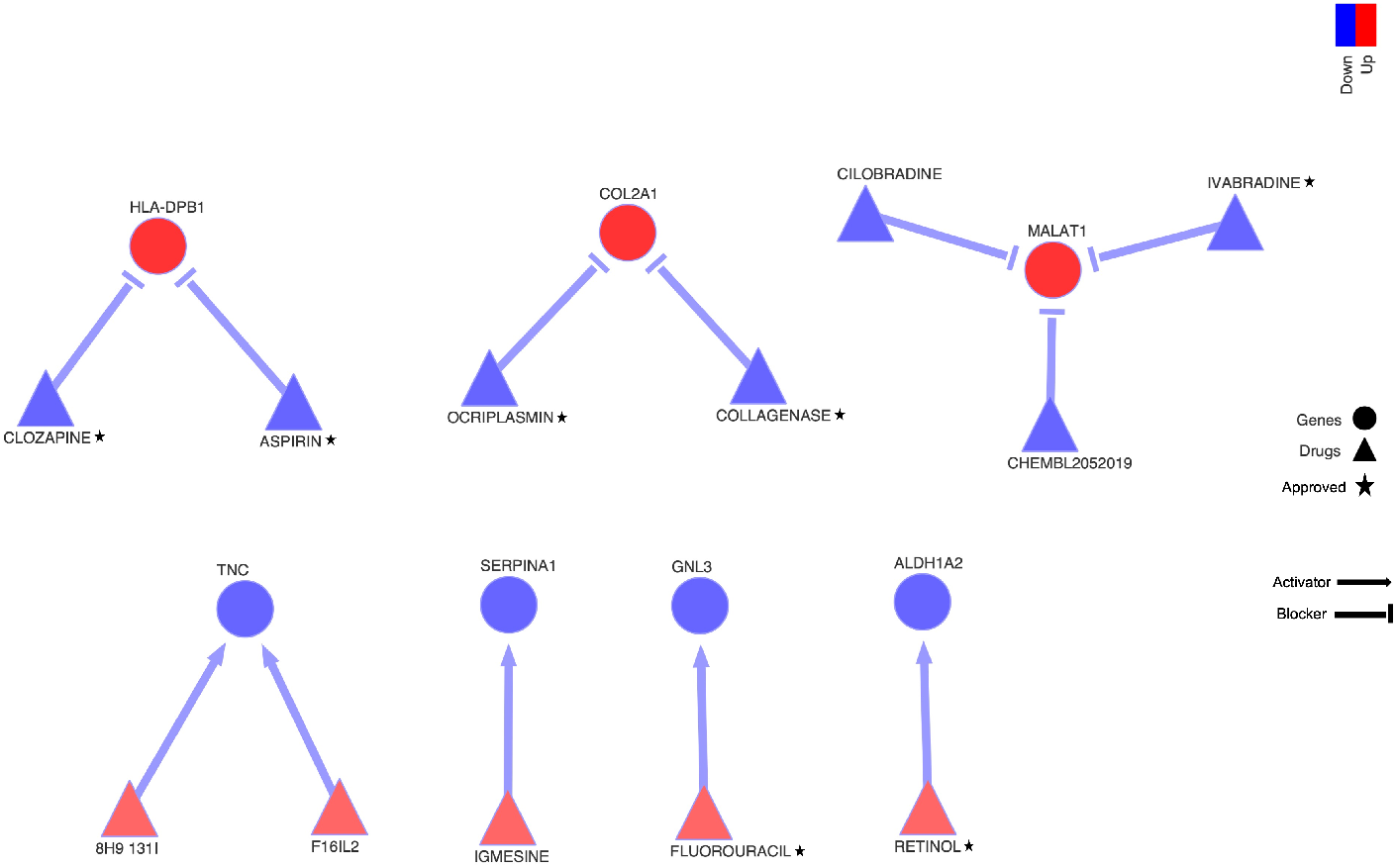
Seven AEI genes interacting with 14 drugs. Genes are represented by circles and drugs are triangles. The start shows drug approved federal agencies according to the database. Blue are genes with AEI downregulated effect and for drugs represents the blockers interaction. Red are genes with AEI upregulated effect and drugs with an activator interaction.

## Discussion

In the current manuscript, we outlined an unbiased approach to identify positional proxy transcript SNPs of currently strong OA risk loci by AEI analyses in transcriptome-wide datasets of the OA relevant tissues articular cartilage and subchondral bone from OA patients. Furthermore, we integrated drug-gene interaction database to identify potential drugs that could counteract effects the OA risk alleles in the relevant tissue. Subsequently, we identified 10 genes subject to AEI in cartilage (**Table 2**), that were not previously recognized as the main gene by GWAS, including two lincRNAs *MALAT1* and *ILF3-DT*. Moreover, we identified AEI in HLA-DPA1/DPB1 and MGP genes also in subchondral bone. We advocate that these genes should be prioritized for further, translation towards underlying biological mechanisms e.g. in human 3D in vitro model systems incorporating bone and/or cartilage tissue units.

To our knowledge, we are the first to identify AEI of lincRNAs *MALAT1* and *ILF3-DT* in articular cartilage, highlighting that the OA risk is associated to upregulated expression (meta-φ=0.54 and meta-φ=0.6, respectively, **Table 2**). Although these two lincRNAs were also expressed in subchondral bone (data not shown) we could not find significant effect due to the low number of heterozygous in this dataset. The *Metastasis Associated Lung Adenocarcinoma Transcript 1* (*MALAT1*) is a well-conserved lncRNA involved in several diseases, and it is known to play a role in transcription, epigenetic regulation and splicing(14). *MALAT1* regulate inflammation, cellular responses to oxidative stress and cellular senescence(15). Noteworthy, *MALAT1* is decreased in senescent cells(16) and in bleomycin-induced murine fibrosis where myeloid deletion of MALAT1 increases susceptibility to fibrosis and the number of profibrotic M2 macrophages(17). MALAT1 was identified as as sponges for miR-150-5p and it was found up-regulated in OA chondrocytes compared with normal chondrocytes(18). Nevertheless, MALAT1 was not significant differentially expressed between lesioned and preserved cartilage(19) or in subchondral bone(20) in our dataset. We found allelic imbalance of the allele C (in LD with the risk allele G) with an upregulated effect (**Figure 2B**). It is tempting to speculate that *MALAT1* risk allele could be partially regulating inflammation, or even senescence pathways in OA. Being highly tissue specific and condition-specific expression pattern lincRNAs have been proposed as new tool for a particular and personalized therapeutic approach(21). In addition, cartilage is a postmitotic tissue, highly dependent on adequate epigenetic mechanisms to establish dynamic changes in gene expression, which make both lincRNAs (*MALAT1* and *ILF3-DT)* promising candidates to be use as therapeutical tools. Nevertheless, more function studies (e.g CRISPR-cas9) on these regulatory variants will be necessary to clarify this issue.

Even though we took a different approach than our previously allelic transcriptomic-wide study (10), we replicated previously reported AEI in the important OA risk genes, *MGP, TNC* and *WWP2* in cartilage. Additionally, we showed AEI for *MGP* in both cartilage and subchondral bone tissues with the same direction of effect but different SNPs in high LD with the top GWAS SNP (downregulation effect of the rs1800801-T risk allele, meta-φ=0.37 in cartilage and rs4236-C risk allele meta-φ=0.33 in subchondral bone, **Table 3**). Previously *MGP* was identified with a low expression level for the risk allele in cartilage, fat pad and synovium, however in blood *MGP* expression was increased (8).Here we confirmed similar effect for AEI in *MGP* in the two main OA related tissues, cartilage and subchondral bone. In this regard, *TNC* and *WWP2* appeared to be showing AEI specifically in articular cartilage. Despite the fact that they are robustly expressed in subchondral bone and had a reasonable number of heterozygote samples (seven and three, respectively) they did not show significant AEI in this tissue. Henceforth, these OA risk alleles may likely confer risk to OA particularly via aberrant function in articular cartilage. Nevertheless, further confirmation in a larger transcriptomic dataset of subchondral bone is necessary to further corroborate this statement.

A notable finding presented here is the highly significant and consistent AEI of the OA risk gene *TNC* in articular cartilage. *TNC* encodes the glycoprotein Tenacin that is abundantly expressed (75^th^ percentile) in the extracellular matrix (22). TNC is a key molecule in cartilage remodelling and participates in chondrogenesis and cartilage development (23). *TNC* may be involved in mechanotransduction in response to mechanical stress (22). The here reported AEI indicated that the OA risk allele T (rs2274836 SNP) is associated to downregulation of *TNC* in cartilage of OA patients, but *TNC* was upregulated in lesioned as compared to preserved OA cartilage. The latter suggesting a beneficial *TNC* response of chondrocytes in cartilage during the OA pathophysiology e.g. to restore tissue homeostasis. These findings are in line with *in vitro* cells models (24) showing that *TNC* expression levels was still normal at the early OA phase, but with its expression disappearing at the late phase with cartilage maturation. In addition, it has been shown that administration of intra-articular TNC in rabbits using scaffolding matrices, promoted the repair of cartilage defects (25), suggesting, that *TNC* deficiency enhance cartilage degeneration. Together these data indicate that aberrant *TNC* signaling may confer important risk to OA, as such should be prioritized for functional follow up research towards underlying OA disease mechanisms.

*COLGALT2* was among the nine genes with differential expression between preserved and lesioned cartilage (meta-φ=0.54 and FC=1.42, **Figure 3B**; **Table 2**). *COLGALT2* encodes procollagen galactosyltransferase 2, an enzyme that post-translationally glycosylates collagen. This gene has been also previously identified with methylation quantitative trait loci (mQTL) (26). Based on the mQTL it was speculated that the expression changes in this gene is mediated by differential enhancer methylation (26). The authors further suggested that a reparative response by *COLGAT2*. Recently, using several molecular biology tools, including CRISPR-Cas9, Kehayova et al.,(5) proposed that the OA risk allele from SNP rs11583641, mediates decreased levels of DNA methylation in cartilage at the COLGALT2 enhancer, which ultimately increased expression of the gene (5). Our data support these findings, since we showed the SNP rs12023991 in high LD with the most associated GWAS SNP rs11583641, with upregulation effect of the risk allele (meta-φ=0.54, FDR = 0.00132), followed by an upregulation of *COLGALT2* in lesioned cartilage (FC=1.42, FDR =1.92×10^−5^), suggesting allele-specific co-regulation element that could explain these differences. The upregulation could require methylation however only via the risk allele.

The prioritization of druggable targets based on genetic support has been proven to highly increase efficacy of clinical trials. OA GWAS genes can result in candidate targets for drug discovery (7). We integrated allelic expression imbalance of OA risk variants (or variants in LD with it) with a large drug-gene target database (DGBIv4). We were able to identify 14 drugs interacting with at least one AEI gene (**Figure 4**). Of note, SERPINA2 gene interacts with Igmesine drug, which could potentially reverse the allelic downregulation effect by enhancing SERPINA2 expression. Nevertheless, this drug has not yet being approved by federal agencies to be use, therefore more studies to confirm this candidate should be performed. In the other hand, we found seven drugs which has been already approved to be used, including Retinol which interacts with *ALDH1A2* gene and Ivabradine which interacts with the lncRNA MALAT1. The Retinol binding protein 4 (RBP4) acts as an immunomodulatory adipocytokine (a bioactive product produced by adipose tissue), and recently was found positively correlated with MMP-1 and MMP-3 in chondrocytes and highly expressed in synovial fluid and plasma from OA patients(27), suggesting a possible role of this molecule in the OA pathogenesis. Moreover, Ivabradine was found to induce up-regulation of matrix metalloproteinase-3 (MMP-3) and MMP-13 at gene and protein levels(28), which are key genes for OA pathophysiology. In addition, as we suggested above, the lncRNA MALAT1 appears to have an important role in OA. MALAT1 in chondrocytes promotes chondrocyte proliferation, suppress chondrocyte apoptosis and reduces extracellular matrix degradation (29,30). Recently, the use of Revesratrol to suppress the activation of MALAT1 showed a decreased of pro-inflammatory genes (NF-κB1 and IL-6) by modulating a microRNA (miR-9) (31). Considering that Ivabradine is a blocker modulator, we can speculate that this drug could act in a similar way as Reverstrol with MATLAT1 to reverse the effects of OA response. Nevertheless, more *in-vitro* and *in-vivo* studies will be essential to clarify this hypothesis.

Taken together, by applying AEI in disease-relevant tissues showed that we can improve the selection of possible causal genes associated to OA. Moreover, we were able to identify new candidate genes OA, including two lincRNAs. This was the first time that lincRNA involved in OA showed a direct interaction with approved drug. Thus, by pinpointing the risk allele which influence gene expression in disease-relevant tissues, we can pave the way for further post-GWAS *in-vitro* models in OA.

## Methods

### Samples

Ethical approval for the Research Arthritis and Articular Cartilage (RAAK) study(32) was obtained from the medical ethics committee of the Leiden University Medical Center (P08.239/P19.013), and informed consent was obtained from all patients included. The current study included 65 OA patients (39 knees and 26 hips) (**Table 1**) who underwent a joint replacement surgery, from which macroscopically unaffected (preserved N=56) cartilage entered the analyses complemented with independent samples for which we had available affected (lesioned N=9) OA articular cartilage only. In addition, unaffected subchondral bone from 26 OA patients all overlapping with cartilage samples were also included.

**Table 1:** Samples characteristics

| **Cartilage** | **Total (N=65)** | **RNAseq - hip**<br>**(N =26 )** | **RNAseq - knee (N=39)** |
| --- | --- | --- | --- |
| Age (SD) | 64.5 (16.7) | 64.2 (16) | 64,6 (17.4) |
| Females | 54 | 20 | 34 |
| **Bone** | **Total (N=26)** | **RNAseq - hip**<br>**(N=6)** | **RNAseq – knee (N=18)** |
| Age (SD) | 68.1 (9.5) | 67.8 (8.8) | 65.7 (8.5) |
| Females | 27 | 6 | 16 |

**Table 2:**
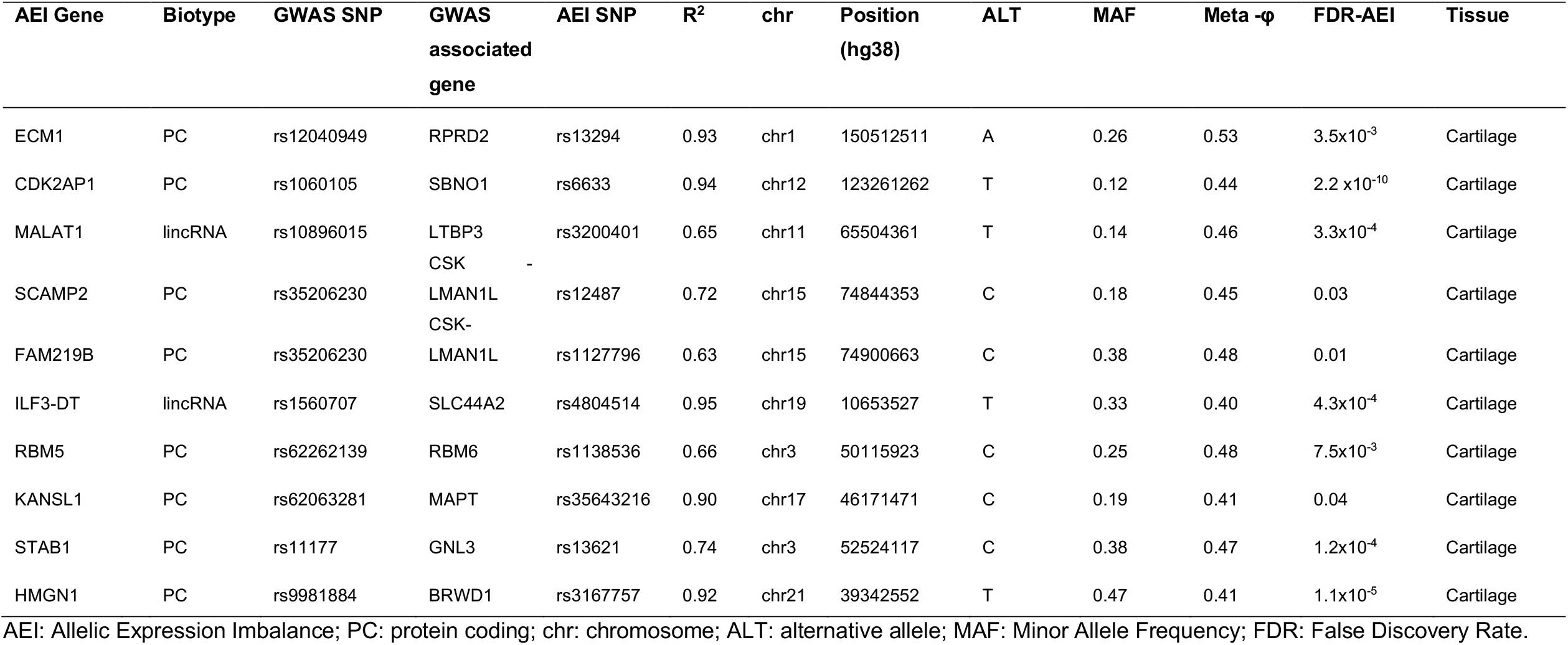
AEI genes not reported in GWAS catalog.

**Table 3:** AEI genes differentially expressed in cartilage and bone.

| AEI<br>DEG | Average Expression<br>(Normalized read counts) | Fold<br>Change | FDR<br>DEG | GWAS<br>SNP | AEI<br>ALLELE | r<sup>2</sup> | Meta -ϕ | FDR<br>AEI |
| --- | --- | --- | --- | --- | --- | --- | --- | --- |
| *COLGALT2* | 1918.57 (Upper quantile-75%)<br>(cartilage) | 1.42 | 1.92x10<sup>-5</sup> | rs11583641_C | rs12023991_C | 1 | 0.46 | 0.021 |
| *ILF3-DT* | 142.32 (Upper quantile-75%)<br>(cartilage) | 0.69 | 3.41x10<sup>-4</sup> | rs1560707_T | rs4804514_T | 0.95 | 0.4 | 4.3 x10<sup>-4</sup> |
| *MGP*<br>(bone) | 38663.18 (Upper quantile-75%)<br>(cartilage)<br>(3826.17- (Upper quantile-75%)) | 1.43<br>(1.53) | 0.02<br>(NS) | rs4764133_T | rs1800801_T | 0.95 | 0.37<br>(0.33) | 1.06x10<sup>-6</sup><br>(1.1x10<sup>-19</sup>) |
| *RFLNA* | 90.98 (Down quantile-50%) | 0.48 | 6.48x10<sup>-5</sup> | rs4765540_C | rs12551_C | 0.74 | 0.48 | 0.006 |
| *SLC44A2*<br>(bone) | 1014.102 (Upper 75%)<br>(1383.285(Upper quantile-75%)) | 0.80<br>(0.84) | 0.028<br>(0.019) | rs1560707_T | rs1053007_G | 0.95 | 0.34<br>(NS) | 1.6x10<sup>-9</sup><br>(NS) |
| *SMAD3* | 492.69 (Upper quantile-75%) | 0.83 | 0.028 | rs12901071_A | rs1061427_A | 0.60 | 0.45 | 0.01 |
| *TNC* | 21349.84 (Upper quantile-75%) | 1.41 | 0.010 | rs2480930_A | rs2274836_T | 1 | 0.47 | 2.08x10<sup>-7</sup> |
| *WWP2* | 5381.64 (Upper quantile 75%) | 0.79 | 0.034 | rs34195470_A | rs2270841_C | 0.65 | 0.54 | 0.0046 |
| *CHTS3* | 2509.1042 (Upper quantile 75%) | 1.23 | 2.9x10<sup>-2</sup> | rs3740129_A | rs4148950_A | 0.8 | 0.41 | 2.8x10<sup>-7</sup> |

AEI: Allelic Expression Imbalance; DEG: Differentially Expression Gene; FDR: False Discovery Rate; NS: Non-significant. Fold change direction related to the lesioned tissue.

### RNA-Sequencing

Total RNA from articular cartilage and subchondral bone was isolated using Qiagen RNeasy Mini Kit (Qiagen, GmbH, Hilden, Germany). Paired-end 2×100 bp RNA-Sequencing (Illumina TruSeq RNA Library Prep Kit, Illumina HiSeq2000 and Illumina HiSeq4000) was performed. Strand specific RNA-seq libraries were generated which yielded a mean of 20 million reads per sample. Detailed description for quality control (QC) alignment, mapping and normalization are described earlier for cartilage (11) and for subchondral bone (12). In addition, common population SNP sites from the Genome of the Netherlands (33) were masked during read alignment to prevent potential reference alignment bias. AEI events were assessed on SNPs called using SNVMix2 with default settings (34) with minimum coverage of 25 and at least 10 reads per allele. Raw autosomal SNP-level data, for SNPs with ≥ 10 reads, was annotated by assigning heterozygous SNPs to genes using Ensembl v97 and SNPdb v151.

### Allelic Expression Imbalance

To perform AEI, we used transcriptome-wide sequencing (RNA-seq) data after QC from articular cartilage (N=65) and subchondral bone (N=24) from OA patients. While the cartilage samples were not fully paired (56 preserved and 9 lesioned cartilage), subchondral bone samples were paired (24 pairs) and overlapping with cartilage. First, we calculated the LD of each 100 associated loci reported on GWAS catalog database(13) (**Supplementary Table 3**), taking a window of 1Mb from the most associated variant reported by each study. To identify AEI events, we used our previously transcriptome-wide approach(10). First, the count fraction of the alternative alleles among the alternative and reference alleles together (φ) was determined for each heterozygous individual. Finally, a meta-analysis was performed to generate a meta-φ and P-value for each genetic variant with a false discovery rate (FDR) correction for multiple tests. Additionally, differential expression analysis between paired samples of preserved and lesioned cartilage (N=35 pairs) and subchondral bone (N=24) was verified in our previous published datasets (11,12), and further intersected with all genes reported to be associated to OA in the GWAS catalog database. We assess the average normalized expression in cartilage and subchondral bone using the DESeq2 normalization approach (35) for all OA associated genes.

### Data integration with Drug–Gene Interaction Database (DGIdb 4.0)

The Drug Gene Interaction Database (DGIdb v4.0) (36) was used to determine potentially druggable targets for all genes that show allelic expression imbalance. Currently, the DGIdb database contains more than 40,000 genes (including protein-coding genes and non-coding RNAs) and 10,000 drugs involved in over 15,000 drug-gene interactions. To identify drugs known to act directly on the specific gene we limited the DGIdb interaction group (IG) score >0.50. This score is based on: (1) the amount of evidence from different sources supporting the drug-gene (i.e. the number of publications and different databases); (2) the number of genes set on each search that interact with the given drug; (3) the degree to which the drug has known interactions with other gene (36). Moreover, to identify enrichment of the overlapping AEI genes with genes interacting with drugs in the DGI database, we applied a Fisher’s exact test using all genes expressed in articular cartilage and subchondral bone (N= 21,666) as background. Finally, to prioritize drug-gene targets in OA context we removed drugs showing a mode of action inconsistent with the AEI direction.

## Data Availability

Data are available upon reasonable request by any qualified researchers who engage in rigorous, independent scientific research, and will be provided following review and approval of a research proposal and Statistical Analysis Plan (SAP) and execution of a Data Sharing Agreement (DSA). All other data relevant to the study are included in the article.

https://ega-archive.org/studies/EGAS00001004476

## Acknowledgements

We thank all study participants of the RAAK study. The Leiden University Medical Centre have and are supporting the RAAK. We thank Evelyn Houtman, Marcella Hoolwerff, Enrike van der Linden, Robert van der Wal, Peter van Schie, Shaho Hasan, Maartje Meijer, Daisy Latijnhouwers, Anika Rabeling-Hoogenstraaten, and Geert Spierenburg for their contribution to the collection of the joint tissue. The research leading to these results has received funding from:

- TreatOA which is funded by the European Commission framework 7 programme grant 200800.
- BBMRI Metabolomics Consortium funded by BBMRI-NL, a research infrastructure financed by the Dutch government (NWO, grant nr 184.021.007 and 184033111).
- Dutch Scientific Research council NWO /ZonMW VICI scheme (91816631/528)

## Data availability statement

RNA-seq data are deposited at the European Genome-Phenome Archive (accession number: EGAS00001004476) and ArrayExpress (E-MTAB-7313). Data are available upon reasonable request by any qualified researchers who engage in rigorous, independent scientific research, and will be provided following review and approval of a research proposal and Statistical Analysis Plan (SAP) and execution of a Data Sharing Agreement (DSA). All other data relevant to the study are included in the article

## List of Supplementary Tables

Supplementary Table 1:Overview of allelic expression imbalance SNPs in cartilage and bone.

Supplementary Table 2:Drug interaction with allelic imbalance expression genes.

Supplementary Table 3:GWAS catalog results for osteoarthritis used in this study.

## Abbreviations

OA: Osteoarthritis
GWAS: Genome-wide association study
AEI: Allelic expression imbalance
LD: Linkage disequilibrium
*COLGAT2*: Collagen Beta(1-O)Galactosyltransferase 2
MGP: Matrix Gla Protein
SNPs: Single nucleotide polymorphisms
LincRNA: Long intergenic non-coding RNAs
*MALAT1:*: Metastasis Associated Lung Adenocarcinoma Transcript 1
FDR: False Discovery Rate
*ILF3-DT*: Interleukin Enhancer Binding Factor 3 Divergent Transcript
HLA: Human Leukocyte Antigen
*HLA-DPA1:*: Human Leukocyte Antigen-heterodimer consisting of an alpha
*HLA-DPB1:*: Human Leukocyte Antigen-heterodimer consisting of an alpha
*TNC:*: *Tenascin C*
FC: Fold Change
*SLC44A2:*: Solute Carrier Family 44 Member 2
DGBI: Drug Gene Interaction Database
IG: Interaction group
CRISPR-cas9: Clustered Regularly Interspaced Short Palindromic Repeats Associated Protein 9
*WWP2*: *WW Domain Containing E3 Ubiquitin Protein Ligase 2*
mQTL: methylation Quantitative Trait Loci
SERPINA2: Serpin Family A Member 2
*ALDH1A2:*: *Aldehyde Dehydrogenase 1 Family Member A2*
RBP4: Retinol binding protein 4
MMP-3: matrix metalloproteinase-3
MMP-13: matrix metalloproteinase-13
NF-κB1: Nuclear Factor kappa B1
IL-6: Interleukin-6
miR-9: microRNA-9

